# Plasma glial fibrillary acidic protein is an early marker of Aβ pathology in Alzheimer’s disease

**DOI:** 10.1101/2021.04.11.21255152

**Authors:** Joana B. Pereira, Shorena Janelidze, Ruben Smith, Niklas Mattsson-Carlgren, Sebastian Palmqvist, Henrik Zetterberg, Erik Stomrud, Nicholas J. Ashton, Kaj Blennow, Oskar Hansson

## Abstract

Although recent clinical trials targeting amyloid-β (Aβ) in Alzheimer’s disease (AD) have shown promising results, there is increasing evidence suggesting that understanding alternative disease pathways that interact with Aβ metabolism and amyloid pathology might be important to halt the clinical deterioration. In particular, there is evidence supporting a critical role of astroglial activation and astrocytosis in AD. However, to this date, no studies have assessed whether astrocytosis is independently related to Aβ or tau pathology, respectively, *in vivo*. To address this question, we determined the levels of the astrocytic marker glial fibrillary acidic protein (GFAP) in plasma and cerebrospinal fluid (CSF) of 217 Aβ-negative cognitively unimpaired individuals, 71 Aβ-positive cognitively unimpaired individuals, 78 Aβ-positive cognitively impaired individuals, 63 Aβ-negative cognitively impaired individuals and 75 patients with a non-AD neurodegenerative disorder from the Swedish BioFINDER-2 study. Subjects underwent longitudinal Aβ (^18^F-flutemetamol) and tau (^18^F-RO948) positron emission tomography (PET) as well as cognitive testing. We found that plasma GFAP concentration was significantly increased in all Aβ-positive groups compared with subjects without Aβ pathology (p<0.01). In addition, there were significant associations between plasma GFAP with higher Aβ-PET signal in all Aβ-positive groups, but also in cognitively normal individuals with normal Aβ values (p<0.001), which remained significant after controlling for tau-PET signal. Furthermore, plasma GFAP could predict Aβ-PET positivity with an area under the curve of 0.76, which was greater than the performance achieved by CSF GFAP (0.69) and other glial markers (CSF YKL-40: 0.64, sTREM2: 0.71). Although correlations were also observed between tau-PET and plasma GFAP, these were no longer significant after controlling for Aβ-PET. In contrast to plasma GFAP, CSF GFAP concentration was significantly increased in non-AD patients compared to other groups (p<0.05) and correlated with Aβ-PET only in Aβ-positive cognitively impaired individuals (p=0.005). Finally, plasma GFAP was associated with both longitudinal Aβ-PET and cognitive decline, and mediated the effect of Aβ-PET on tau-PET burden, suggesting that astrocytosis secondary to Aβ aggregation might promote tau accumulation. Altogether, these findings indicate that plasma GFAP is an early marker associated with brain Aβ pathology but not tau aggregation, even in cognitively normal individuals with a normal Aβ status. This suggests that plasma GFAP should be incorporated in current hypothetical models of AD pathogenesis and be used as a non-invasive and accessible tool to detect early astrocytosis secondary to Aβ pathology.

## Introduction

There is increasing evidence suggesting that the pathogenesis of Alzheimer’s disease (AD) is not restricted to amyloid-β (Aβ) plaques and tau tangles but also includes strong interactions with immunological mechanisms.^1^ In line with this, astrocyte reactivity or astrocytosis is a well-known pathological process that is commonly found surrounding Aβ plaques in the brains of AD patients.^2^ Although the exact role of astrocytosis is not clear, several studies have shown that reactive astrocytes penetrate Aβ plaques with their processes, possibly in an attempt to isolate the plaques from the surrounding neuropil and phagocytize them.^3,4^ This close relationship between astrocytes and plaques has been further supported by neuropathological reports showing that reactive astrocytes follow the same spatial distribution of Aβ plaques in the association cortex of AD patients.^5,6^

In contrast to the association between astrocytosis and Aβ plaques, the relationship between reactive astrocytes and tau tangles has been less investigated. The few available studies that have assessed this showed that reactive astrocytes also interact with tau, but only in advanced stages of AD by penetrating the extracellular ghost tau tangles.^7,8^ In addition, it has also been found that the association between reactive astrocytes and tau tangles parallels the progression of AD,^9^ however it is not clear whether this association is independent of Aβ plaques, which are normally present in the brains of patients with AD who have tau tangles.

Thanks to the development of biomarkers to measure astrocytosis, such as the glial fibrillary acidic protein (GFAP), it is now possible to address this question and disentangle the underlying effects of reactive astrocytes on both Aβ and tau pathology *in vivo*. In particular, the recent assays that allow measuring the concentrations of GFAP in the blood have already demonstrated the potential of plasma GFAP in distinguishing different stages of AD and detecting Aβ positivity on positron emission tomography (PET).^10-18^ However, no study has yet assessed whether plasma GFAP is also associated with tau-PET burden. Moreover, to this date, it is not known whether astrocytosis is independently related to Aβ or tau pathology, respectively, *in vivo*. This is important for several reasons including the fact that Aβ and tau pathology are not independent processes, showing a synergistic and complex interaction over the course of AD that may become exacerbated in the presence of other pathological processes such as astrocytosis.^19^ Thus, a better understanding of when reactive astrocytes emerge in the progression of AD and how they relate to the classic AD pathologies is crucial to determine their clinical value as diagnostic or prognostic tools as well as to inform the development of anti-inflammatory drugs in clinical trials.

The aim of this study was to assess whether plasma and cerebrospinal fluid (CSF) GFAP concentrations change across different stages of AD and investigate the independent relationships between these markers with Aβ and tau pathology measured using ^18^F-flutemetamol PET and ^18^F-RO948 PET, respectively. Moreover, we compared the performance of GFAP to other glial markers such as soluble triggering receptor expressed on myeloid cells 2 (sTREM2) and chitinase 3-like 1 (YKL-40) to detect Aβ and tau-PET pathology in order to determine the specificity of our findings. We also evaluated the prognostic ability of baseline GFAP markers to predict longitudinal changes in PET burden and cognitive decline. Finally, we conducted sensitivity analyses in a separate group of patients with non-AD disorders to assess the ability of GFAP markers to detect Aβ-positivity determined with CSF Aβ42/40.

## Materials and methods Participants

This study included 504 individuals from the Swedish BioFINDER-2 cohort (NCT03174938), a prospective study with the aim of developing new biomarkers for the early diagnosis of AD and other neurodegenerative disorders. All participants were recruited at Skåne University Hospital and the Hospital of Ängelholm in Sweden between 2017 and 2020 and included cognitively unimpaired controls, patients with mild cognitive impairment, AD dementia and non-AD disorders.^20^ Further details regarding the inclusion and exclusion criteria of the subjects included in the different BioFINDER-2 cohorts can be found in the Supplementary Material.

For the purposes of this study, only subjects with baseline plasma and CSF levels of GFAP in addition to other glial markers (YKL-40, sTREM2), ^18^F-flutemetamol PET, ^18^F-RO948 PET and mini-mental examination scores (MMSE) were included. In addition, a subsample (n = 196) also underwent longitudinal PET imaging and cognitive assessments (n = 185). Finally, a group of patients with non-AD neurodegenerative disorders without ^18^F-flutemetamol PET data was also included.

### Plasma and cerebrospinal fluid biomarkers

Plasma and CSF samples were collected in the morning during the same visit, with participants nonfasting.^20^ Blood was collected in six EDTA-plasma tubes and centrifuged (2,000 g, + 4 ° (C) for 10 min. Following centrifugation, plasma was aliquoted into 1.5-ml polypropylene tubes (1 ml plasma in each tube) and stored at −80 °C within 30–60 min of collection. CSF was collected by lumbar puncture and stored at −80°C in polypropylene tubes.^18^ The following assays were used to measure the different biomarkers of interest to this study: GFAP Discovery kits for HD-X (Quanterix®, Billerica, MA, USA) for plasma GFAP; NeuroToolKit robust prototype assays (Roche Diagnostics) for CSF GFAP, YKL-40 and sTREM2; and Meso Scale Discovery immunoassays (MSD; Rockville, MD, USA) for CSF Aβ42 and CSF Aβ40.^20,21^ In all subjects, Aβ status was established using CSF Aβ42/40 levels with a previously established cutoff of 0.0752 defined with mixture modeling^22^, because CSF Aβ42/40 was available in all cases (by study design), whereas Aβ-PET was only available in non-demented cases.^18^

### Imaging acquisition and preprocessing

All subjects underwent ^18^F-flutemetamol PET and ^18^F-RO948 PET on General Electrics Discovery MI scanners. ^18^F-flutemetamol PET images were acquired 90 to 110 minutes after injection of 185 MBq 18F-flutemetamol and ^18^F-RO948 PET images were acquired 70 to 90 minutes after injection of 370 MBq ^18^F-RO948. Images were reconstructed using VPFX-S (ordered subset expectation maximization combined with corrections for time-of-flight and point spread function).

All ^18^F-flutemetamol and ^18^F-RO948 PET images were motion-corrected, time-averaged and coregistered to their corresponding skull stripped, longitudinally preprocessed T1-weighted images. ^18^F-Flutemetamol scans were normalized using a reference region that included the whole cerebellum, brain stem and eroded subcortical white matter,^23^ whereas ^18^F-RO948 images were normalized by a reference region consisting of the inferior cerebellar gray matter.^24^

For ^18^F-flutemetamol images, we calculated the standardized uptake value ratios (SUVR) for a global composite region that included the caudal anterior cingulate, frontal, lateral parietal and lateral temporal gyri.^23^ In contrast, for ^18^F-RO948 PET images we calculated the SUVRs for three composite regions that corresponded to Braak stages I-II (entorhinal cortex), III-IV (parahippocampal, fusiform, amygdala, inferior temporal, middle temporal) and V-VI (posterior cingulate, caudal anterior cingulate, rostral anterior cingulate, precuneus, inferior parietal, superior parietal, insula, supramarginal, lingual, superior temporal, medial orbitofrontal, rostral middle frontal, lateral orbitofrontal, caudal middle frontal, superior frontal, lateral occipital, precentral gyrus, postcentral gyrus and paracentral gyrus).^25^ Finally, voxel-wise analyses were conducted using the preprocessed ^18^F-flutemetamol and ^18^F-RO948 PET images using the statistical parametric mapping software SPM12 (https://www.fil.ion.ucl.ac.uk/spm/) after applying a smoothing kernel of 8 mm.

### Statistical analyses

Logarithmic or reciprocal transformations were applied to the variables that were not normally distributed. Then, a set of pairwise t-tests was used to compare plasma and CSF GFAP levels between Aβ-negative cognitively unimpaired (CU), Aβ-positive CU, Aβ-positive cognitively impaired (CI), Aβ-negative CI and non-AD patients, while adjusting for age and sex, and using CSF Aβ42/Aβ40 to determine the Aβ status.

To assess the ability of CSF and plasma GFAP markers to predict Aβ and tau pathology, we used two different approaches: one based on regions of interest (ROI) and the other one based on whole brain voxel-wise analyses. For the first approach, we built separate linear regression models with plasma or CSF GFAP as the dependent variable and global Aβ, tau stages I-II, tau stages III-IV or tau stages V-VI SUVRs as the outcome, controlling for age and sex. In all models, we verified that the residuals were normally distributed, there was no heteroscedasticity and no multicollinearity. For the second approach, we conducted voxel-wise regression analyses using plasma or CSF GFAP as the dependent variable and the smoothed preprocessed ^18^F-Flutemetamol or ^18^F-RO948 PET images as the outcome, including age and sex as covariates. All ROI-based and voxel-wise analyses were carried out in Aβ-negative CU, Aβ-positive CU, all CU subjects, Aβ-positive CI and Aβ-negative CI individuals.

For the PET variables showing a significant relationship with plasma and CSF GFAP, we performed three additional analyses. First, we built spline models^26^ to determine the trajectories of GFAP markers as a function of higher PET burden over the course of AD. Due to previous evidence showing that AD progresses from Aβ-negative CU to Aβ-positive CU and finally Aβ-positive CI,^27^ we only included these groups in this analysis. Secondly, to determine the impact of astrocytosis on the relationship between the two classical AD hallmarks, we conducted mediation analyses to test whether the relationship between Aβ-PET and tau-PET burden could be explained by a mediation of GFAP, while controlling for age and sex. The significance of the mediation was assessed by calculating bias-corrected 95% confidence intervals (CIs) using bootstrapping (500 resamples). Thirdly, to establish the area under the curve, sensitivity, specific and accuracy of GFAP markers in determining an Aβ-positive status, we calculated receiver-operating curves using a bootstrap procedure with 1000 permutations in the groups who had ^18^F-flutemetamol PET data: all CU subjects, all CI subjects and in the whole sample. Since non-AD patients did not have ^18^F-flutemetamol scans, the analyses in this group were conducted using CSF Aβ42/40 to determine Aβ-positivity. All receiver-operating curve analyses included YKL-40 and sTREM2 to assess the value of GFAP with respect to other glial markers and their performance was compared using the DeLong test.

Finally, to test whether plasma and CSF GFAP markers were associated with longitudinal changes in cognition and PET burden we applied linear mixed effect models. These models used longitudinal MMSE scores or PET SUVRs as a dependent variable and the GFAP markers, time, age, sex as fixed effects. We also included the interaction between biomarker levels and time (together with the main effects), and random effects for intercepts. Separate models were built for plasma and CSF GFAP, which were ran in all CU subjects, all CI individuals and in the whole sample. The models ran across the entire sample included Aβ and cognitive status as additional covariates. In addition, to assess whether the effects of GFAP on cognition were independent of Aβ-PET, longitudinal changes in Aβ-PET burden were also included as a covariate in a secondary analysis.

Statistical analyses were carried out using SPSS 25.0 (IBM Corp., Armonk, NY, USA), R (version 3.5.1) or SPM12. The analyses conducted in R and SPSS were adjusted for multiple comparisons using false discovery rate (FDR) corrections (q < 0.05, two-tailed).^28^ Similarly, the voxel-wise analyses using PET images were adjusted for multiple comparisons with topological FDR corrections in SPM12 (p < 0.05, two-tailed).^29^

## Data availability

Anonymized data will be shared by request from a qualified academic investigator for the sole purpose of replicating procedures and results presented in the article and as long as data transfer is in agreement with EU legislation on the general data protection regulation and decisions by the Ethical Review Board of Sweden and Region Skåne, which should be regulated in a material transfer agreement.

## Results

### Study participants

In total, 504 participants were included in this study, of which 217 were Aβ-negative CU, 71 were Aβ-positive CU, 78 were Aβ-positive CI (mild cognitive impairment or AD dementia), 63 were Aβ-negative CI (mild cognitive impairment) and 75 had a non-AD neurodegenerative disorder. There was a moderate correlation between plasma GFAP and CSF GFAP in the entire sample (r = 0.582, p < 0.001).

### Plasma GFAP concentration is increased across different AD stages, whereas CSF GFAP concentration is increased in non-AD disorders

Our findings revealed that plasma GFAP concentration was lowest in Aβ-negative CI, followed by Aβ-negative CU, Aβ-positive CU and Aβ-positive CI individuals. The group comparisons showed that both Aβ-positive CU and CI individuals showed elevated plasma GFAP levels compared to the Aβ-negative CU group (F_(2,286)_ = 8.33, p = 0.004, F_(2,293)_ = 12.90, p < 0.001, respectively) and to the Aβ-negative CI group (F_(2,132)_ = 18.06, p < 0.001; F_(2,139)_ = 24.14, p < 0.001, respectively). These findings suggest that plasma GFAP is elevated in individuals with Aβ pathology.

In contrast to plasma GFAP, CSF GFAP was significantly elevated in patients with a non-AD disorder compared to both Aβ-negative CU individuals (F_(2,290)_ = 9.61, p = 0.002) and Aβ-positive CU individuals (F_(2,144)_ = 5.71, p = 0.018). These findings suggest that CSF GFAP might be sensitive to different underlying pathological processes unrelated to AD and might be better suited for the detection of non-AD disorders.

### Plasma and CSF GFAP concentrations are associated with Aβ-PET independently of tau-PET burden

To assess whether GFAP biomarkers were associated with the severity of Aβ deposition, we built linear regression models using global Aβ-PET SUVR values as the outcome in addition to plasma or CSF GFAP as the predictors. We found that increasing GFAP levels in plasma were associated with greater global Aβ-PET burden in all CU individuals (t = 4.24, p < 0.001), Aβ-negative CU individuals (t = 2.31, p = 0.022), Aβ-positive CU individuals (t = 2.11, p = 0.039) and Aβ-positive CI individuals (t = 2.88, p = 0.005). These results were further confirmed by the voxel-wise analyses, which showed that plasma GFAP correlated with higher Aβ-PET deposition in several neocortical regions in all CU individuals as well as in Aβ-positive CU individuals. In contrast to plasma GFAP, CSF GFAP only showed a significant association with higher Aβ-PET deposition in Aβ-positive CI subjects (t = 2.92, p = 0.005). To determine whether these results were independent of tau pathology, we repeated all of the above analyses including tau-PET SUVR values of different Braak stage ROIs as covariates. These analyses showed that Aβ-PET burden was still related to increased plasma GFAP in the same groups (all CU individuals: t = 4.50, p < 0.001; Aβ-negative CU individuals: t = 2.70, p = 0.007; Aβ-positive CU individuals: t = 2.32, p = 0.024; Aβ-positive CI individuals: t = 2.08, p = 0.041) and to increased CSF GFAP in Aβ-positive CI individuals (t = 2.18, p = 0.032), after controlling for tau-PET.

To assess whether the GFAP biomarkers were associated with the severity of tau deposition, we built linear regression models using the tau-PET SUVR values across different Braak stage ROIs as the outcome in addition to plasma or CSF GFAP as the predictors. We found that increasing plasma GFAP levels were only associated with tau-PET burden across Braak stages III-IV and V-VI in Aβ-positive CI individuals (III-IV: t = 2.86, p = 0.006; V-VI: t = 2.97, p = 0.004). However, these associations lost their significance after including global Aβ-PET as a covariate (III-IV: t = 1.60, p = 0.114; V-VI: t = 2.00, p = 0.050), indicating that they were not independent of Aβ pathology.

### Plasma GFAP concentration shows early increases with Aβ-PET burden

To determine the trajectories of plasma and CSF GFAP concentrations over the course of AD, we fitted spline models for these markers using global Aβ-PET SUVR as a proxy for time. These analyses did not include non-AD patients or Aβ-negative CI individuals since they are not considered to be part of the AD spectrum.^27^ The results of these analyses revealed that both models were significant, with the one having plasma GFAP as a predictor showing a better model fit (r2: 0.21, p < 0.001) compared to the one with CSF GFAP (r2: 0.14, p < 0.001). The trajectories of these models revealed initial increases for both plasma GFAP and CSF GFAP, which continued rising after reaching the threshold for Aβ-PET positivity and then later came to a plateau. Despite showing similar trajectories, when we compared the splines of plasma and CSF GFAP, we could observe that plasma GFAP showed steeper initial increases, overcoming CSF GFAP levels even before Aβ-PET positivity. These results indicate that plasma GFAP might exhibit greater changes with increasing Aβ pathology during AD compared to CSF GFAP.

### Plasma GFAP may partially mediate the relationship between Aβ-PET and tau-PET over the course of AD

To further investigate the role of astrocytosis in relation to the classical AD pathologies over the course of the disease we conducted mediation analyses in all CU subjects as well as Aβ-positive CI individuals. These analyses showed that plasma GFAP mediated the effect between global Aβ-PET and both tau-PET stages I-II (−0.027, 95% CI: 0.00 to −0.056, P = 0.035) and stages III-IV (−0.027, 95% CI: −0.056 to −0.01, P = 0.010) in CU individuals. Moreover, plasma GFAP also mediated the effects between global Aβ-PET and tau-PET stages V-VI (0.131, 95% CI: 0.00 to 0.33, P = 0.046) in CI individuals. These findings suggest that astrocytosis secondary to Aβ accumulation might be one the factors contributing to tau accumulation in AD.

### Plasma GFAP identifies an Aβ-positive status more accurately than CSF GFAP and other glial markers

To determine the performance of the glial biomarkers (GFAP, YKL-40 and sTREM2) to detect Aβ positivity, we conducted receiver-operating curve analyses in the whole sample, all CU individuals, all CI individuals as well as non-AD patients, regardless of their Aβ status, which were defined using Aβ-PET or CSF Aβ42/40 (non-AD patients only). These analyses showed that plasma GFAP showed an area under the curve of 0.761 in the whole sample, 0.754 in all CU individuals, 0.779 in all CI individuals and 0.755 in non-AD patients. These performances were significantly better than the ones achieved by CSF GFAP (Z = 2.68, p = 0.007), CSF sTREM2 (Z =3.31, p < 0.001) and almost CSF YKL-40 (Z = 1.77, p = 0.077) in the whole sample; by CSF GFAP (Z = 2.24, p = 0.024) in CU individuals; by CSF GFAP (Z = 2.24, p = 0.025), CSF sTREM2 (Z =3.08, p = 0.002) and CSF YKL-40 (Z = 2.72, p = 0.006) in CI individuals; and by CSF YKL-40 (Z = 2.42, p = 0.016) and almost CSF sTREM2 (Z = 1.80, p = 0.072) in non-AD patients. Altogether, these analyses indicate that plasma GFAP can detect an Aβ positive status more accurately than the other glial markers.

### Plasma GFAP concentration is associated with longitudinal Aβ accumulation determined by PET

For the subsample that underwent longitudinal Aβ-PET (n = 196, number of visits: M = 1.5, IQR = 1; follow-up time: M = 1.7 years, IQR = 0.3), we used linear mixed models to evaluate whether the GFAP markers were also associated with Aβ changes over time in a neocortical composite region. These analyses showed that plasma GFAP predicted longitudinal Aβ deposition in the whole sample (t = 2.888, p = 0.004), whereas CSF GFAP did not (t = 1.478, p = 0.141), after controlling for age, sex, baseline Aβ status and presence of cognitive impairment. Notably, these analyses remained significant after adjusting also for tau-PET burden (t = 2.905, p = 0.004), indicating they were independent of tau pathology. However, no significant results when the analyses were conducted in the CU and CI groups separately.

### Plasma and CSF GFAP concentrations are associated with longitudinal cognitive decline

For the subsample that underwent longitudinal cognitive assessment (n = 185, number of visits: M = 2, IQR = 2; follow-up time: M = 1.9 years, IQR = 0.7), we also used linear mixed models to evaluate whether the GFAP markers were associated with cognitive changes over time. These analyses showed that both plasma GFAP and CSF GFAP predicted cognitive decline in the whole cohort, even after adjusting for longitudinal Aβ-PET changes (plasma GFAP: t = −3.303, p = 0.001; CSF GFAP: t = −2.485, p = 0.014). These results suggest that, in addition to being a marker of Aβ pathology, astrocytosis could have an independent negative impact on longitudinal cognition. No significant results were found when the analyses were conducted in the CU and CI groups separately.

## Discussion

Although emerging evidence suggests that inflammation has a causal role in AD,^11-18^ the detection of inflammatory markers has not yet been established as a valuable method for the early diagnosis and monitoring of AD patients.^1^ Our findings show that plasma GFAP holds great potential as an early and specific marker of Aβ deposition even during the earliest stages of AD. Moreover, we found that plasma GFAP was a prognostic marker of both longitudinal Aβ accumulation and cognitive decline and a mediator of the effects of Aβ-PET on tau-PET burden. In light of the invasiveness of lumbar punctures for CSF and the high cost of PET imaging, our findings suggest that plasma GFAP could become a widely available screening tool to identify astrocytosis in early AD. Moreover, it could also be used to evaluate the effects of anti-Aβ therapies on glial activation as well as to better understand the role of astrocytosis over the course of AD.

Activated glia in the form of reactive astrocytes is one of the most prominent neuropathological features of AD, being normally found surrounding Aβ plaques in postmortem brain tissue.^30^ The role of these reactive astrocytes has been debated over the past few years, with some studies suggesting they are part of an endogenous defensive mechanism to eliminate the plaques, whereas others defend that their persistent activation induces a toxic inflammatory process that contributes to worsening AD progression.^31^ Regardless of their role, several studies have used biomarkers such as GFAP to measure astrocytosis *in vivo* in the CSF or more recently in the blood plasma. Although studies assessing GFAP in CSF have reported somewhat inconsistent findings across different stages of AD,^32,33^ recent studies on plasma GFAP showed more promising results. In particular, elevated plasma GFAP was recently found in subjects with subjective cognitive decline, mild cognitive impairment and AD dementia with a positive Aβ-PET scan.^15^ In addition, an association between longitudinal plasma GFAP and conversion to dementia was also found in a prospective clinical cohort, highlighting its potential value as a prognostic tool.^34^ In the current study, we extend these previous findings by showing that plasma GFAP is not only elevated in subjects with Aβ pathology but also correlates with continuous Aβ-PET values, even in individuals with normal CSF Aβ42/40 levels. Interestingly, when we conducted our analyses at the voxel-level we found that elevated plasma GFAP was associated with higher Aβ-PET burden in neocortical regions where Aβ accumulation is normally observed in AD both in all CU as well as Aβ-positive CU individuals. This result is in agreement with previous studies showing that astrocytes show dynamic changes over the course of AD, with reactive astrocytes being more prominent in earlier disease stages.^31^ In addition, this result is also in line with a previous report showing that reactive astrocytes follow the same spatial distribution of Aβ plaques in the association cortex of AD patients.^5^

Compared to Aβ plaques, the potential associations between reactive astrocytes and neurofibrillary tangles have been much less studied. Immunohistochemical and electron microscopy studies have shown that reactive astrocytes can also penetrate with their processes the extracellular ghost tangles in the midst of the neuropil in advanced AD.^7,8^ Moreover, in another study, a significant linear rise of astrocytosis in the vicinity of neurofibrillary tangles was found with increasing AD progression, although this relationship was weaker than the one observed between astrocytosis and Aβ plaques.^35^ Finally, in a study using autoradiography, tau deposits were observed in similar brain areas as activated astrocytes, supporting a pathological interconnection.^36^ To our knowledge, no studies have assessed whether astrocytic markers such as plasma and CSF GFAP are associated with *in vivo regional* tau-PET pathology in the course of AD. Here, we show that greater tau-PET signal in middle and late Braak stages was associated with increasing plasma GFAP in Aβ-positive CI individuals. However, when these associations were adjusted for Aβ-PET burden their significance was lost, in contrast to the correlations between plasma GFAP and Aβ-PET, which remained unchanged after adjusting for tau-PET signal. These findings indicate that astrocytosis measured by initial increases in GFAP is specifically associated with Aβ pathology, challenging previous assumptions that both Aβ and tau pathology can trigger reactive astrocytes in AD.^6,35^

In contrast to plasma GFAP, CSF GFAP was only associated with Aβ pathology in CI individuals, showed less steep increases with increasing Aβ-PET burden in the spline models, and was significantly elevated in patients with non-AD disorders. These findings suggest that plasma and CSF GFAP might be measuring partially different pathological processes, with the former being more closely related to abnormal Aβ accumulation due to AD, whereas the latter also incorporating other neuroinflammatory changes unrelated to Aβ pathology. Moreover, together with sTREM2 and YKL-40, CSF GFAP showed a lower sensitivity and specificity in detecting Aβ-positivity compared to plasma GFAP. In fact, the ability of plasma GFAP to identify Aβ-PET or CSF Aβ42/40 pathology was quite consistent across the whole sample, CU individuals, CI individuals and non-AD disorders the with an area under the curve above 0.75 in all cases, which is similar to the values obtained in a previous study using the same biomarker.^15^ These results suggest that plasma GFAP can be reliably used to detect Aβ-positivity across different disease stages as well as non-AD disorders. The differences between plasma and CSF GFAP could also be related to different clearance pathways of the molecule into the biofluids. For instance, astrocytic end-feet projections to the neurovascular unit may provide a direct clearance pathway of the molecule into the bloodstream, and there may also be relationships with vascular amyloid pathology, which is common in AD^37^.

Regarding the prognostic value of GFAP, we found that both CSF and plasma predicted global cognitive decline to the same degree, even after adjusting for changes in Aβ-PET accumulation. It has been previously suggested that, although glial responses are initially triggered by Aβ burden, they can become progressively independent of Aβ with disease progression and contribute to neurodegenerative and cognitive changes.^35^ Our findings seem to confirm this assumption as the associations between plasma GFAP and cognitive decline were still significant after adjusting for Aβ-PET burden, indicating that astrocytosis has a negative impact on cognition that goes beyond its link to Aβ pathology. This suggests that astrocytosis may play a role in promoting cognitive deterioration in AD and that therapies targeting this neuroinflammatory process could at least partially ameliorate cognitive symptoms. In addition, regarding longitudinal Aβ accumulation, we found that plasma GFAP was a better predictor of Aβ-PET burden over time, even after controlling for tau-PET signals, suggesting that plasma GFAP is a more sensitive tool to identify not only baseline but also future Aβ pathology. Finally, our mediation analyses revealed that plasma GFAP also mediated the effects between Aβ-PET and tau-PET in CU and CI individuals in a stage-dependent manner, indicating that astrocytosis might be contributing to tau accumulation, although this effect is not independent of Aβ pathology. These findings could have important implications for current treatments in AD and suggest that a combination of anti-amyloid therapies with anti-inflammatory treatments could potentially reduce the formation of tau aggregates.

Our study has several strengths, including the large number of participants with several glial biomarkers and longitudinal Aβ-PET, longitudinal tau-PET and global cognition. However, a few limitations should also be recognized such as the fact that we did not have serial longitudinal measures of plasma and CSF GFAP, which would have been useful to determine their trajectories over the course of AD and determine their potential clinical value as longitudinal monitoring tools. In addition, plasma GFAP and CSF GFAP were also measured using different platforms, Simoa and NeuroToolKit, respectively. At the time of the study, there was only one follow-up available for Aβ-PET and two follow-ups available for cognition within a period of approximately two years. This may have limited our ability to detect effects that were stronger enough to reach significance in the CU and CI groups separately since a longer time period may be required to observe more prominent Aβ-PET and cognitive changes. Finally, the inclusion of a PET imaging tracer such as ^11^C-deuterium-l-deprenyl^38^ would have been interesting to include to assess the relationship between plasma GFAP and regional brain astrocytosis.

In summary, here we show that baseline plasma GFAP seems to be a very early marker of astrocytosis associated with Aβ pathology suggesting it can be used to detect baseline Aβ-positivity and predict future Aβ accumulation and cognitive decline. In addition, contrary to previous assumptions, astrocytosis measured with GFAP was not associated with tau pathology after controlling for Aβ, indicating it is not only an early but also a quite specific marker for Aβ pathology. Although current models of AD have adopted a neurocentric view that starts with Aβ accumulation, followed by tau deposition and neurodegeneration, it is well known that neurons cannot function properly without the proper support of glial cells such as astrocytes.^31^ Thus, our findings highlight the importance of including astroglial markers in the cascade of pathological changes occurring in AD, particularly plasma GFAP, which could potentially be used as a non-invasive tool to evaluate the effects of anti-Aβ drugs or anti-inflammatory treatments on astrocytosis in clinical trials.

## Data Availability

Anonymized data will be shared by request from a qualified academic investigator for the sole purpose of replicating procedures and results presented in the article and as long as data transfer is in agreement with EU legislation on the general data protection regulation and decisions by the Ethical Review Board of Sweden and Region Skane, which should be regulated in a material transfer agreement.

